# Novel Application of a Multistate Model to Evaluate the Opioid Use Disorder Care Cascade: A Retrospective Cohort Study

**DOI:** 10.1101/2022.03.10.22271924

**Authors:** Anarina L. Murillo, Tianyu Sun, Hilary Aroke, Jeffrey Bratberg, Stephen Kogut, Brandon D.L. Marshall, Jesse L. Yedinak, Josiah D. Rich, Rebecca Lebeau, Joseph W. Hogan, Ashley Buchanan

## Abstract

**Background:** Evaluating the opioid use disorder (OUD) care cascade can improve OUD treatment retention and care.

**Objectives:** To identify risk and protective factors for retention among patients in OUD treatment.

**Methods:** We conducted a retrospective cohort study among patients diagnosed with OUD using data from the Rhode Island (RI) All-Payer Claims Database from 2011 to 2019. Patients who initiated treatment (Stage 2) were classified into sub-stages of retention (Stage 3) corresponding to multistate modeling states capturing early retention (sub-stage 1), short and long-term retention (sub-stage 2), and short and long-term disengagement (sub-stage 3). The association of baseline characteristics with state transitions were evaluated.

**Results:** A cohort of 6,939 RI residents diagnosed with OUD included 41% aged 40 to 60 years, 57.6% male, and 70.8% Medicaid beneficiaries. In sub-stage 1, cannabis (Relative risk ratios (RRR) = 1.16; 95% confidence interval (CI) = 1.04,1.29) and cocaine use disorders (RRR=1.15; 95% CI=1.05,1.25) increased early disengagement risk after engagement. Medicaid beneficiaries were less likely to experience early disengagement (RRR=0.81; 95% CI =0.76,0.87). In sub-stage 2, alcohol (RRR=1.29; 95% CI=1.13,1.47) or cocaine use disorders (RRR=1.18; 95% CI=1.01,1.40) increased risk of disengagement among patients in the retention states. In sub-stage 3, tobacco (RRR=1.10; 95% CI=1.01,1.21) and alcohol (RRR=1.14; 95% CI=1.03,1.27) use disorders were associated with re-engagement from disengaged states.

**Conclusion:** The multistate model applied to a cohort of patients initiating medication for OUD led to the identification of factors associated with treatment engagement and retention. These results may guide strategies to sustain treatment among OUD patients.

## Introduction

Extra-medical opioid use and opioid use disorder (OUD) has been identified as a national public health crisis by the US Department of Health and Human Services (1, 2). While medications for OUD—including methadone, buprenorphine, and naltrexone—has helped to curtail the opioid overdose crisis in the United States (US) (1-3), less than 20% of people with OUD received medication for OUD in the US in 2019 (3). Thus, there is an urgent need to expand the availability of OUD treatment, promote equity in access, ensure uptake, and achieve a sustained reduction in overdose (4, 5).

Several barriers to access and uptake of OUD treatment contribute to low utilization of these medications, including provider, pharmacy, geographic, and regulatory factors, as well as financial barriers (4-7). Prior to the COVID-19 pandemic, patients with OUD were required to receive their methadone in an office-based setting only or were prescribed buprenorphine after an in-person consultation, and prescribers were required to complete training and apply for a DEA waiver to prescribe buprenorphine (8, 9). Some changes have been made during the COVID-19 pandemic to improve access to OUD care, including take-home medication for methadone and buprenorphine initiation via telemedicine (9, 10). However, despite these policy changes, medication for OUD treatment gaps persist (11, 12).

An improved understanding of the OUD care cascade informs more effective interventions and policy for treatment initiation and retention (11, 13-16). The OUD care cascade is typically defined using the following stages: diagnosed with OUD (Stage 1); initiate medications for OUD (Stage 2); retention in treatment (Stage 3); and recovery (Stage 4) (14). Previously reported OUD care cascade studies frequently rely on limited cross-sectional, patient-level data and often involves macro-level summaries, risk factor analysis, or simulation techniques (11, 14, 17). However, patients at risk and with OUD can receive intermittent care, rendering their engagement in care as a time-varying process. Cross-sectional analyses have known limitations, such as a lack of temporal ordering, and offer only a partial picture of the OUD care cascade (11, 14, 17).

In this study, we developed and applied a multistate model to study the OUD care cascade longitudinally. By studying longitudinal patient trajectories, we can identify protective and risk factors that inform intervention strategies for particular stages of OUD care (18, 19). We applied a multistate model to the Rhode Island All-Payer Claims Database (RI APCD) to examine risk and protective factors among patients initiated in treatment for OUD (Stage 2) and retained in therapy (Stage 3) using the OUD care cascade as a framework. Hence, through a novel application of a multistate model to the RI APCD, our study gives insight into factors associated with transition along the OUD care cascade among patients diagnosed with OUD in RI, including comorbidities and other clinical diagnoses, such as hepatitis C virus and HIV diagnoses.

## Methods

### Data Source and Study Population

Deidentified data for all individuals enrolled in the RI APCD from January 1, 2011 to Dec 31, 2019 were used in this study. The RI APCD is a large-scale database that routinely collects healthcare medical and prescription claims data from a variety of payer sources, including Medicare, RI Medicaid, and RI’s nine largest commercial payers (20), capturing up to 80% of RI residents (20). The enrollment files contain demographic data and dates of enrollment.

This database undergoes three steps in validation and quality assurance checks and 95% of the commercial claims are finalized within three months, while 80 to 85% of the Medicaid claims are finalized within the same time frame. The prescription drug file contains the record of all prescriptions reimbursed by any of the insurers included in the RI APCD, and includes the therapeutic class code, national drug codes, and date of prescription fill, quantity dispensed, days of supply, as well as pharmacy and prescriber information. The study protocol was reviewed and approved by the Brown University Institutional Review Board.

Our study sample included individuals with an OUD diagnosis in their medical claims data using ICD-9-CM and ICD-10-CM classification codes (21) for opioid abuse or opioid dependence diagnoses (see Appendix A Tables 1-6). A flow diagram describing the sample selection process is in Appendix A Figure 1. Patients with an inpatient admission with OUD as primary or secondary diagnosis, an emergency department visit with OUD as primary or secondary diagnosis, or an outpatient visit with OUD as primary diagnosis were classified as having OUD. We counted the first OUD claim in the database during the study period as their initial OUD diagnosis. Patients aged 18 years or older at the time of their OUD diagnosis with at least 30 days of follow-up after their OUD diagnosis were included; however, we did not exclude individuals who had medication for OUD or related services prior to an OUD diagnosis.

**Table 1.** Baseline^a^ Demographic and Clinical Characteristics of Patients in the Rhode Island All Payers Claims Database, 2011-2019 (N = 6,939).

| Characteristic | Number (%) |
| --- | --- |
| Age at index date |  |
| <40 years | 3602 (52) |
| 40 – 60 years | 2814 (41) |
| 60+ years | 523 (8) |
| Biological Sex |  |
| Female | 2939 (42) |
| Male | 4000 (58) |
| Insurance Type |  |
| Medicaid | 4916 (71) |
| Other (Commercial or Medicare) | 2023 (29) |
| HIV | 135 (2) |
| HCV | 817 (12) |
| Opioid Use Disorder Medication |  |
| Methadone | 850 (12) |
| Buprenorphine | 1483 (21) |
| Naltrexone | 67 (<1) |
| Non-fatal Opioid Overdose | 187 (3) |
| Substance Use Disorder |  |
| Alcohol | 1665 (24) |
| Cannabis | 555 (8) |
| Cocaine | 902 (13) |
| Tobacco | 2220 (32) |
| Pain |  |
| Low back | 1734 (25) |
| Abdominal | 1804 (26) |
| Neck | 69 (<1) |
| Behavioral Disorders |  |
| Anxiety | 3538 (51) |
| Mood | 3816 (55) |
| Personality Disorder | 346 (5) |

<sup>a</sup>The baseline period was defined as 365 days before the first OUD diagnosis in the database.

**Figure 1:**
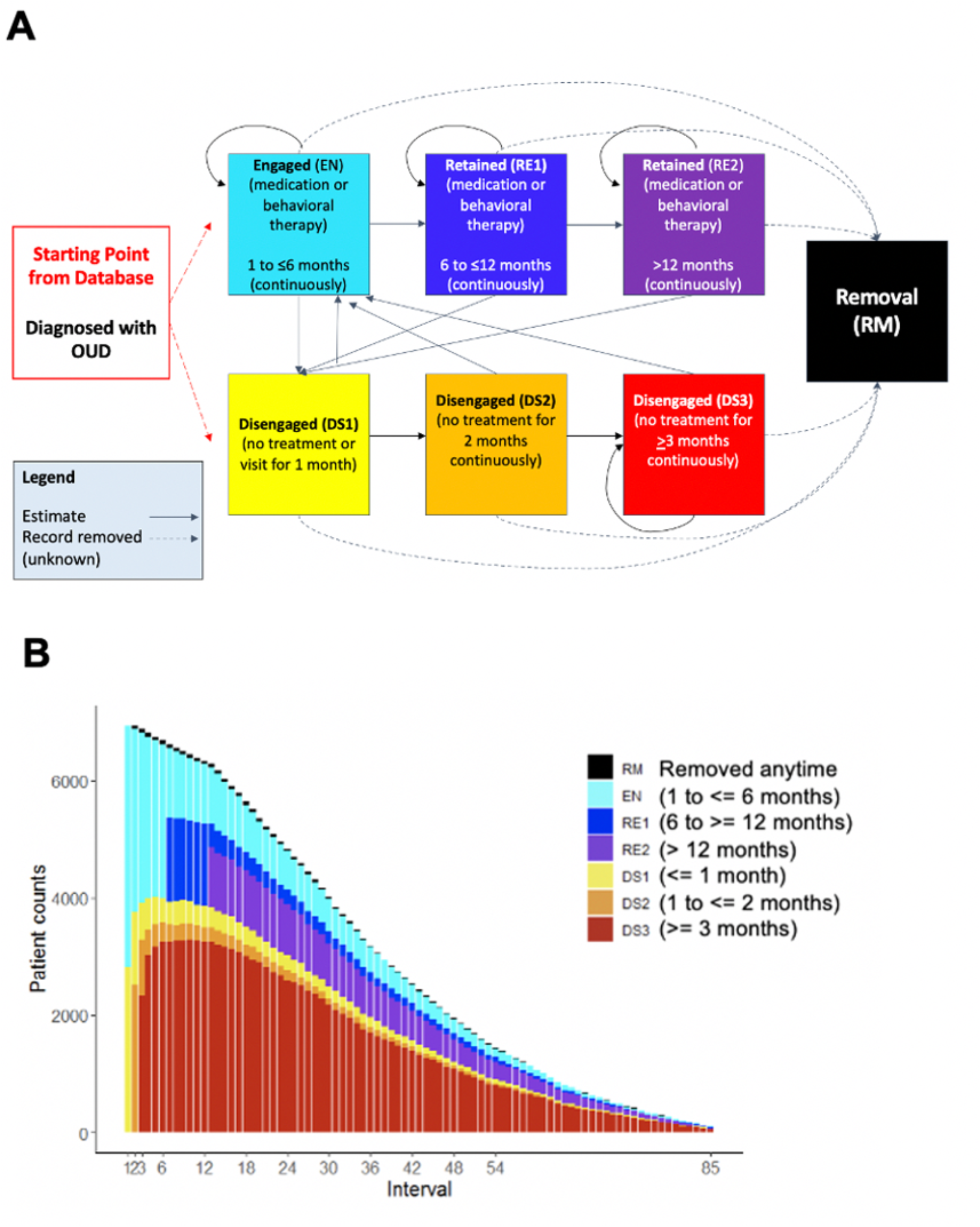
Flow diagram describing the transition states in the Rhode Island OUD Care Cascade for patients identified as receiving both behavior and/or medication therapy (panel A). The total number of patients in each state during the study period (panel B).

Patients needed to be continuously enrolled in the RI APCD with a gap of less than 15 days for disenrollment, enrolled for at least one year before their initial OUD diagnosis in the database, and enrolled no later than December 2018 to allow at least one calendar year of follow-up before administrative censoring. Patients with more than one enrollment record for a given time period had their information combined into a single patient record for that period. Follow-up for patients ended at the time of administrative censoring on December 31, 2019, or at the time of patient disenrollment. The baseline period was defined as 365 days before the first OUD diagnosis in database. Due to a data use agreement, patients enrolled in Medicare Fee for Service (FFS) were excluded prior to the data extraction for this study.

Medications for OUD were determined from the pharmacy claims data using national drug codes (NDCs) and from the medical data using current procedural terminology codes (CPT) (see Appendix A Tables 4-5). Behavioral therapy was identified from the medical claims using CPT and Healthcare Common Procedure Coding System (HCPCS) Codes (see Appendix A Table 3). Treatment for OUD was defined as receiving at least a seven-day supply of OUD medications, CPT for OUD medications, or behavior therapy during a given interval after OUD diagnosis. This included Buprenorphine (with or without Naloxone), Methadone, and Naltrexone (see Appendix A Tables 4-5) (21).

Other demographic and clinical characteristics that may influence patients’ transition along the OUD care cascade were evaluated in our model at baseline only including: age at index was categorized into patients < 40 years, 40 to 60 years, and > 60 years, biological sex (female versus male), and insurance type according to the first claim (Medicaid versus Commercial or Medicare) (22). Clinical, substance use disorder, and behavioral disorders measured at baseline only were included using ICD-9-CM and ICD-10-CM codes (see Appendix A Tables 6). Clinical variables included: diagnosis of HIV, patients with hepatitis C virus (HCV identified by either diagnosis or medication dispensing) (23), and claims for an opioid overdose for poisoning by opium, methadone, heroin, opiates, and narcotics. Substance use disorder variables defined included substances such as tobacco, cocaine, cannabis, and alcohol. Behavioral disorders and clinical factors included those who had a Charlson Comorbidity Index (CCI) > 1, anxiety, personality disorder, pain (e.g., lower back, neck, headache, fibromyalgia, osteoarthritis, rheumatoid arthritis, sprain), and mood disorder. Psychotropic medications such as benzodiazepines, antidepressants, antipsychotics, stimulants, muscle relaxants, sleep aids, and gabapentin use defined as having one or more prescription claim(s) (24) were included (see Appendix A Tables 1-5).

### Multistate Model

The cyclical nature of OUD treatment engagement and retention is modeled as a time-varying process by which individuals diagnosed with OUD move through six states of the OUD care cascade. Figure 1 displays the six states and possible transitions between these states using an OUD care cascade as a framework (14). Patients with an OUD diagnosis in medical claims enter states of the model where they are eligible to receive either medication or behavioral therapy, or both (Stage 2 of (14)) and then transition into the defined sub-stages of retention (Stage 3 of (14)). Sub-stages of OUD care cascade correspond to states in the multistate model defining early engagement and retention (sub-stage 1), short and long-term retention (sub-stage 2), and short and long-term disengagement (sub-stage 3). Using the patient-level data from the RI APCD, we constructed a longitudinal data set with records representing 30 days; if there was a gap of less than 15 days for engagement, we assumed the patient remained engaged with treatment.

In sub-stage 1, the treatment engagement state (EN) was defined as at least one medical encounter for behavioral therapy, at least one CPT code for methadone, or a seven-day supply of OUD medication within one month of the incident OUD diagnosis. We applied a comprehensive definition of treatment for OUD to be defined as receiving behavioral and/or medication therapy. We presented this as the main analysis because receipt of methadone may not be well captured in the database and would possibly underestimate patient engagement with the OUD care cascade. Methadone clinics typically bill per longer episodes and not by the day, so the days of supply of methadone cannot be accurately determined from the claims. Furthermore, some methadone clinics may be supported by public sector grants or support in RI, which would not be captured in the database due to the provider not submitting an insurance claim. A sensitivity analysis that analyzed medication therapy (M model) only in comparison to the behavior and/or medication therapy (BM model) is presented in Appendix C Figures 7 to 9. Patients in the EN state were defined as individuals engaged in treatment (either one behavioral therapy session or had at least 7 days of medications for OUD supply) within one month (or 30-day interval) from their initial OUD diagnosis. Patients who re-engaged at each subsequent month for six months remained in the EN state until transitioning to the short-term retention state (RE1). Patients who did not receive treatment for an entire month can transition from the EN state to the short-term disengaged state (DS1), at which point, they can re-enter the EN state after receiving treatment.

In sub-stage 2, patients retained on treatment for more than 6 months and up to 12 months were considered to be in the short-term retention state (RE1) and those retained in treatment for longer than 12 months were in the long-term retention state (RE2). In sub-stage 3, patients enter the DS1 by either failing to initiate treatment within one month from OUD diagnosis or have no record of treatment for one month after being in either RE1 or RE2 (shown in Figure 1A). Long-term disengagement states were defined as individuals that had disengaged for at least two (DS2) months or three or more months (DS3). Patients who left the model system through disenrollment or death were considered removed (RM). State definitions are detailed in Appendix D Table 7.

The multistate model characterizes individual-level membership of patients diagnosed with OUD into these discrete states as a function of time: EN, retained in OUD care (RE1, RE2), disengaged from OUD care (DS1, DS2, DS3), and disenrolled or deceased (RM) (described in Appendix B and C). Once in the RM state, patients cannot re-enter any states in the model. The proportions of patients in each state were calculated (see Figure 1B). Patient-level transition probabilities between health states (sub-stages) and the association of baseline individual-level predictors with time-varying transitions between states of the OUD care cascade were evaluated using the multistate model (18, 19). State transitions were modeled as a function of covariates using multinomial regression for repeated measures to evaluate associations between individual-level risk and protective factors and transition between the states. Covariates included demographic and clinical characteristics measured at baseline only. Clinical characteristics associated with each of the states were analyzed using generalized estimating equations with a log link and a binomial distribution (25). Relative risk ratios (RRR) were calculated to identify risk and protective factors for transition between the three sub-stages of the OUD care. To quantify uncertainty in the model, we reported 95% Wald-type confidence intervals (CI) with the standard error calculated using either a robust estimator of the variance or a bootstrap resampling method accounting for within subject correlations (26, 27).

This multistate model relies on several assumptions. This multistate assumes first order Markov dependence (18, 19), which means the current state only depends on the immediately preceding state. This was evaluated by also including state membership at two time steps prior as a categorical covariate. We performed empirical checks for a lack of fit for the multistate model by constructing plots of observed versus fitted state membership probabilities for each time interval. The predicted proportions, or predicted marginal state membership probabilities, were calculated using the multistate model with covariates to estimate predicted state membership probabilities at each time point. We calculated the standard error of the predicted proportion in each state using a bootstrap resampling method with 200 bootstrap replicates (26). Our results (shown in Appendix B Figure 2) indicated that the observed and predicted marginal state probabilities were similar during the study period except for the first days of removed, and short (RE1) and long-term (RE2) retention states. We evaluated the second order dependence comparing the observed and predicted state probabilities (see Appendix B Figure 3) and estimated the relative risk ratios when the multistate model also included the second order state membership (see Appendix B Figure 4 to 6). These results show no indication of a lack of model fit and the results appeared to be fairly robust to the first order Markov assumption (18, 19).

**Figure 2:**
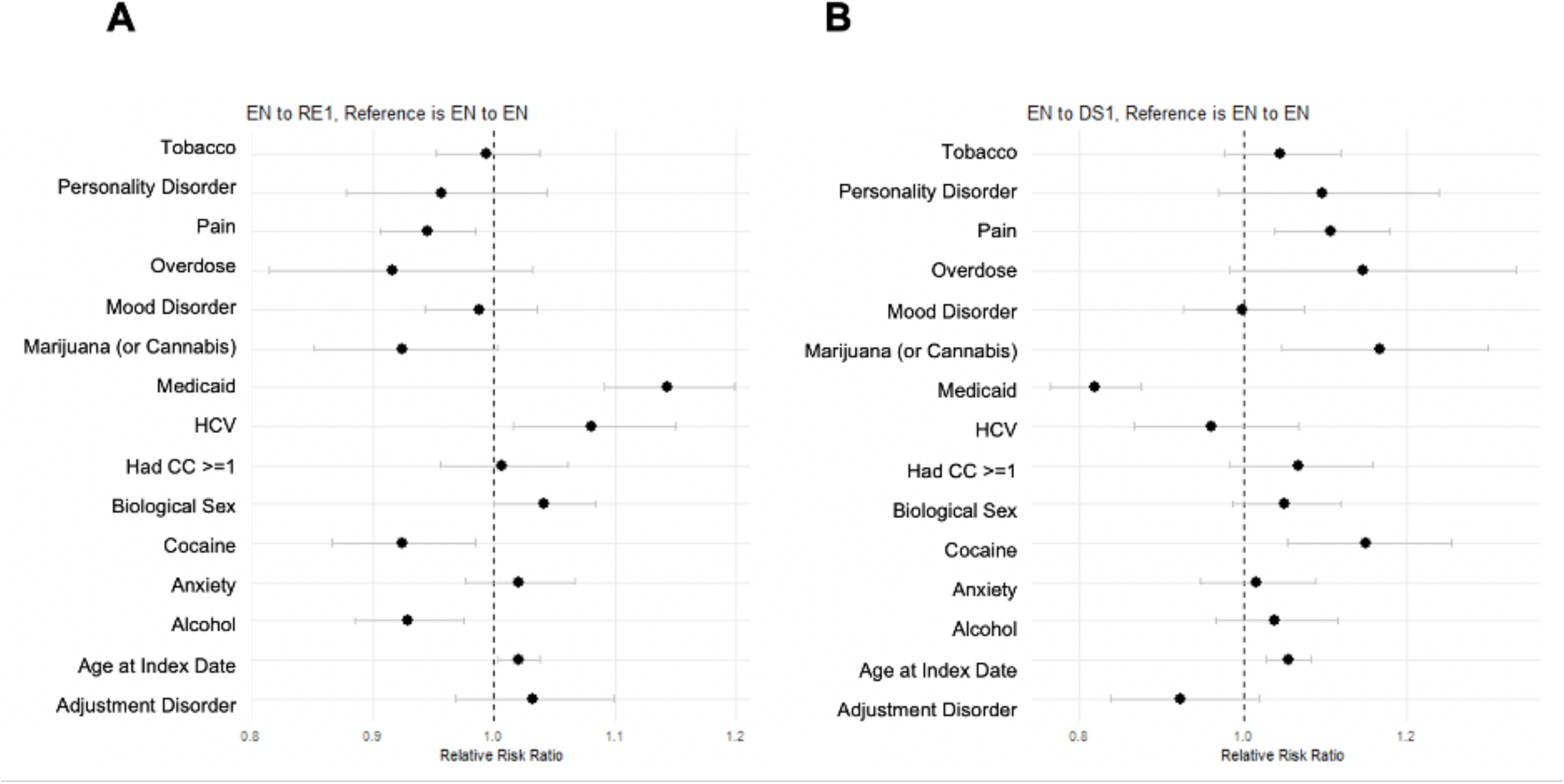
Relative risk ratios^a^ capturing transitions from engagement to early retention (EN to RE1, panel A) and from engagement to early discontinuation (EN to DS1, panel B) among a cohort of patients diagnosed with OUD in Rhode Island, 2011-2019. Patients were treated with behavior and/or medication therapy. ^a^ Reference group for variables defined as follows: “other (commercial or Medicare)” for Medicaid, “male” for biological sex, and “no” for tobacco use disorder, personality disorder, pain, overdose, mood disorder, marijuana (or cannabis), HCV, had CC >= 1, cocaine use disorder, anxiety disorder, alcohol use disorder, and adjustment disorder. Age at index date measured in years.

**Figure 3:**
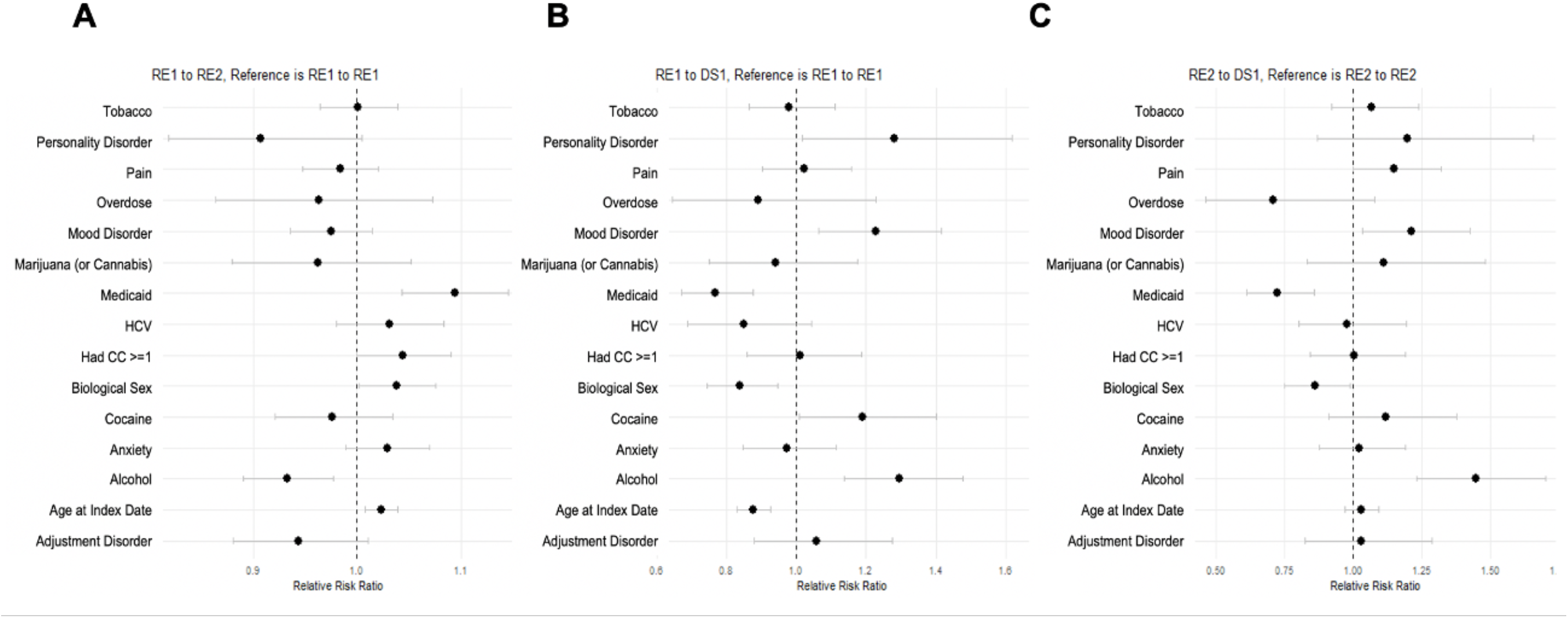
Relative risk ratios^a^ capturing short- and long-term retention. RE1 to RE2 in (panel A), RE1 to DS1 in (panel B), and RE2 to DS1 in (panel C) for patients treated with behavior and/or medication therapy. ^a^ Reference group for variables defined as follows: “other (commercial or Medicare)” for Medicaid, “male” for biological sex, and “no” for tobacco use disorder, personality disorder, pain, overdose, mood disorder, marijuana (or cannabis), HCV, had CC >= 1, cocaine use disorder, anxiety disorder, alcohol use disorder, and adjustment disorder. Age at index date measured in years.

**Figure 4:**
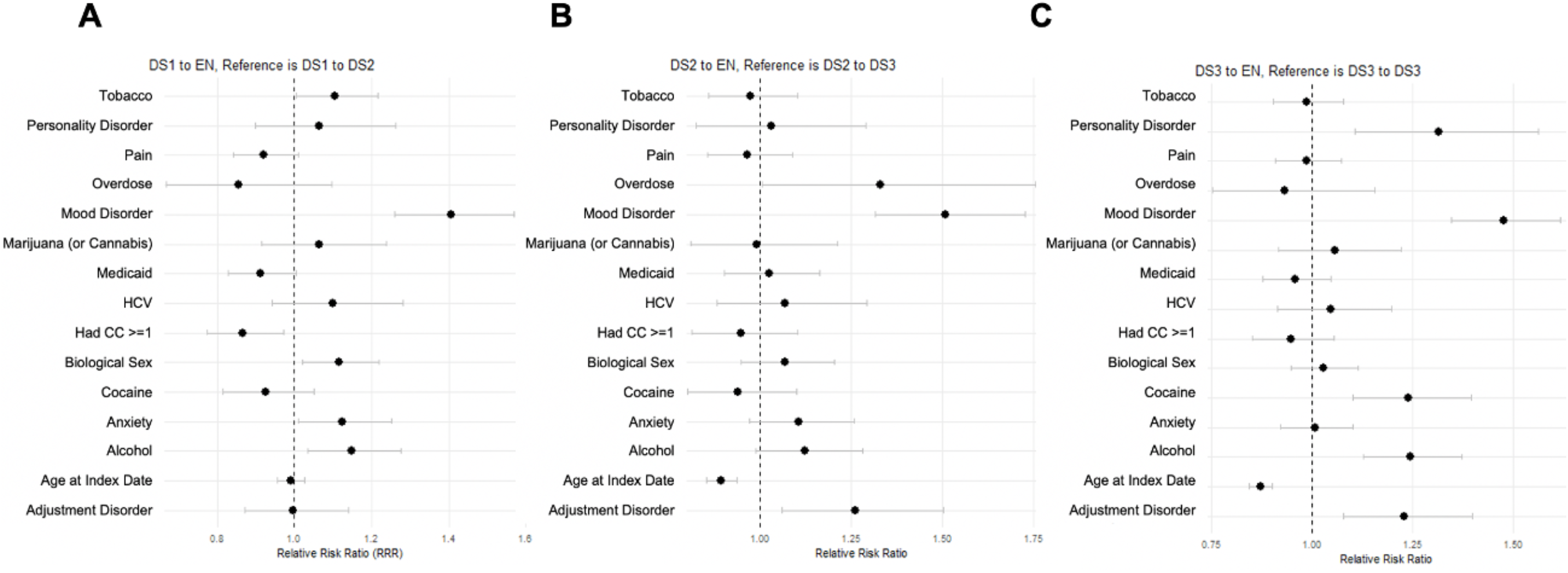
Relative risk ratios^a^ capturing short- and long-term disengagement. DS1 to EN in (panel A), DS2 to EN in (panel B), and DS3 to EN in (panel C) for patients treated with behavior and/or medication therapy. ^a^ Reference group for variables defined as follows: “other (commercial or Medicare)” for Medicaid, “male” for biological sex, and “no” for tobacco use disorder, personality disorder, pain, overdose, mood disorder, marijuana (or cannabis), HCV, had CC >= 1, cocaine use disorder, anxiety disorder, alcohol use disorder, and adjustment disorder. Age at index date measured in years.

## Results

### Clinical Characteristics

The sample included 6,939 adults diagnosed with OUD residing in RI with a majority aged 18 to 39 years (52%) and 58% of patients were male (Table 1). Medicaid was the main insurance source for 71% of patients, whereas Commercial or Medicare were used by the remaining 29% of the sample. Less than 2% of patients were diagnosed with HIV at baseline, and approximately 12% of the sample had documented HCV at baseline. Less than 3% of the population had opioid overdose-related claims and the CCI was greater than 1 for 25% of the sample during the baseline period. Patients claims at baseline for substance use disorder, behavioral disorders, and other clinical conditions are summarized in Table 1.

Patients in the EN state remained engaged at a probability of 0.129 and transition to RE1 with a probability of 0.0150. A patient retained in the RE1 transitions to RE2 with a probability of 0.008 and a probability of 0.121 for remaining in the RE2 state. However, patients identified in the DS3 state remain disengaged with a probability of 0.470. This means that once patients disengage there is almost an average 50% probability that they will remain disengaged during the entire period, while there is only a 2% probability of patients moving onto RE1 from EN. In the following subsections, we summarize the risk and protective factors identified in the three sub-stages (Stages 2 and 3 (14)) in the OUD care cascade: EN and RE1 or DS1 (sub-stage 1), RE1 or RE2 (sub-stage 2), and DS1, DS2, or DS3 (sub-stage 3). State transition probabilities detailed in Appendix D Table 9 and Appendix D Figure 10.

#### Sub-stage 1: Engagement and Early Retention or Disengagement

Several baseline clinical factors were associated with risk of DS1 or RE1 relative to remaining in the EN state (Figure 2A-B). Medicaid insurance beneficiaries (compared to other insurance types) were 19% less likely to experience DS1 (RRR=0.81; 95% CI = 0.76, 0.87). The co-occurrence of other substance use disorder at baseline was associated with a higher risk of transitioning to DS1 from EN including for cannabis (RRR=1.16; 95% CI = [1.04,1.29]) and cocaine (RRR=1.15; 95% CI = [1.05,1.25]). Patients with a pain condition at baseline had a 10% higher risk of DS1 (RRR=1.10; 95% CI = 1.03,1.17) relative to remaining EN. Patients were more likely to experience RE1 from EN with a 2% increase associated with a one year increase in age (RRR = 1.02; 95% CI = 1.00,1.03) and an 8% increase among those with documented HCV, compared to those without HCV (RRR = 1.08; 95% CI = [1.01,1.15]). Medicaid beneficiaries also had a 14% increased likelihood of RE1, compared to those without Medicaid coverage (RRR = 1.14; 95% CI = 1.09, 1.19).

#### Sub-stage 2: Short and Long-Term Retention

Relative to staying in RE1, co-occurrence of alcohol use disorder was associated with a 7% decrease in the likelihood of transitioning to RE2, compared to those without alcohol use disorder (RRR = 0.93; 95% CI = [0.89,0.97]) (see Figure 3A). Medicaid insurance (RRR = 1.09; 95% CI = [1.04,1.14]) was a protective factor increasing the likelihood of RE2 relative to remaining in RE1 and was associated with a 28% reduced risk of DS1 from RE2 (RRR=0.72; 95% CI = 0.61, 0.85). Alcohol (RRR = 1.29; 95% CI = [1.13,1.47]) and cocaine use disorder (RRR = 1.18; 95% CI = [1.01, 1.40]) increased risk of transitioning to DS1 from RE1 (see Figure 3B). Similarly, the co-occurrence of behavioral disorders such as mood disorder increased risk of DS1 from RE1 (RRR = 1.22; 95% CI = [1.06, 1.41]) and from RE2 (RRR = 1.21; 95% CI = [1.03,1.42]). Diagnosed personality disorder was also associated with a 28% increased risk of DS1 from RE1 (RRR=1.28; 95% CI= 1.01,1.61).

#### Sub-stage 3: Re-Engagement after Short and Long-Term Disengagement

Relative to staying in DS1, DS2, or DS3, several protective factors were associated with promoting re-engagement (see Figure 4A to C). The likelihood of transitioning from DS1 to EN, in reference to DS2 was increased by the co-occurrence of tobacco use disorder (RRR=1.10; 95% CI = [1.01,1.21]) and alcohol use disorder (RRR=1.14; 5% CI = [1.03,1.27]). The co-occurrence of alcohol use disorder was associated with higher likelihood of EN from DS3 (RRR=1.24; 95% CI = [1.12,1.37]), relative to remaining in DS3. Re-engagement was higher among patients with diagnosed personality disorder, anxiety disorder, or mood disorder at baseline across the three disengagement states (DS1, DS2, and DS3). Cocaine use disorder (RRR=1.23; 95% CI = [1.10,1.39]) was associated with moving to EN in reference to DS2. Mood disorder at baseline was associated with about a 40% higher likelihood of EN, in reference to DS1, DS2 or DS3 (RRR=1.40; 95% CI = [1.25,1.57]; RRR=1.50; 95% CI = [1.31,1.72]; RRR=1.47; 95% CI = [1.34,1.61], respectively).

## Discussion

We conducted a retrospective cohort study of 6,939 adults diagnosed with OUD residing in RI from 2011 to 2019 using data from the RI APCD. We classified patients in well-defined stages and sub-stages of the OUD care cascade to assess the protective and risk factors for successful transition along the OUD cascade in RI to contribute to the development of interventions to prevent OUD among those at risk, bolster treatment for OUD, and sustain patient recovery in RI and nationwide.

Prior studies of the OUD care cascade have been conducted in Florida (11), RI (14, 17) and British Columbia using routinely-collected administrative data. In Florida (11), the care cascade framework was used to study treatment quality and outcomes of Medicaid beneficiaries in 2017 to 2018; the authors found a reduction in mortality among patients with medication for OUD claims (2%) in comparison to those without (10%). Among those who remained on treatment, there was a 77% reduction in the hazard of mortality (95% CI = 0.17, 0.29), compared to those who disengaged from treatment. In a retrospective cohort study among people with OUD in British Columbia (16), OUD treatment engagement was associated with younger age, male biological sex, substance use disorder, HCV, and homelessness or income assistance. In a recent cross-sectional study using routinely-collected data in RI, about 55% of the at-risk population was diagnosed with OUD and 27% had initiated medication for OUD, while only 18% were retained on treatment in 2016 (14).

In this work, several notable findings on the associations of substance use disorder claims, diagnosis of behavioral disorders, and insurance coverage give insight into possible approaches to increase successful transition along the OUD care cascade. In the early stages, substance use disorders, such as cannabis and cocaine, at baseline increased the risk of short-term disengagement. Similarly, both alcohol and cocaine use disorder, increased the risk of disengagement following short- and long-term retention. This finding is consistent with Macmadu et al. (17) in which 18,374 Medicaid recipients in RI with an opioid overdose or an OUD diagnosis were examined to identify predictors of enrollment in treatment for OUD. Their study concluded that alcohol use disorder or opioid overdose decreased the likelihood of timely engagement in medication-based therapy. These findings and our results suggest that ‘high impact’ settings for OUD screening and referrals to treatment for OUD include programs treating patients for alcohol and other substance use disorders, pain clinics, inpatient psychiatric hospital, and other outpatient behavioral care settings. Our study showed that a patient with substance use disorder promoted re-engagement, thus, protecting against disengagement.

Interestingly, we observed some agreement in risk factors identified by these cross-sectional and longitudinal studies, notably the role of other substance use disorders and pain management in the delivery of OUD care. Patients with comorbidities may experience increased health care engagement as a result of receiving treatment for multiple conditions and, in turn, this may increase opportunities for providers to re-engage patients for OUD care. These results suggest multifaceted approaches to care for patients with OUD that harmonize care, pain management, and polysubstance use disorders (28) in one central facility or through mobile delivery of services to reach patients in their communities (29). We observed an association between Medicaid insurance status and treatment retentions, possibly due to differences in insurance coverage or the quality of services as compared with some commercial payers. Such aspects could not be fully understood from these data.

This study has several important limitations. The definition of OUD was not a previously validated measure and this definition does not capture disease severity; however, we used sets of claims similar to other studies of OUD (11, 14). Although the RI APCD captures many individuals with insurance products, the claims data may not fully capture all adults receiving treatment for OUD. Our study does not include those who disenrolled from these insurance products or those who are incarcerated and possibly enrolled in the RI Department of Corrections medication for OUD treatment program (30-32). Another limitation is that since we counted the first OUD claim in the database during the study period as their initial diagnosis it is possible that patients diagnosed outside of this APCD database were excluded from analysis. Removal in our model includes both death and disenrollment, rendering the bias due to censoring difficult to address analytically due to different mechanisms. Assessing recovery is challenging in claims data because we do not have a discharge status for patients. Some patients enrolled in Medicare FFS may have been censored due to transfer to this insurance product, and for patients with FFS only, they were excluded prior to the data extraction for this study. Lastly, we recognize that behavioral treatment only for diagnosed OUD is not a recommended treatment alone; however, due to the possibly misclassification of methadone treatment in the APCD, we expanded our definition of treatment to protect against underestimating treatment for OUD in the RI APCD. Furthermore, treatment adherence was not assessed, and we cannot assume that filling a prescription for OUD care reflects patient adherence to that medication.

Future work may include linking data from the Department of Corrections and other state and national databases, such as the Prescription Drug Monitoring Program and the National Death Index, to provide a more comprehensive evaluation of the OUD care cascade. New methods are needed in the multistate model to evaluate causal effects in the presence of treatment-confounder feedback to disentangle effects from associations for exposures that can be conceptualized as implementable future interventions (33).

## Data Availability

All data produced in the present study are available upon reasonable request to Brown University.

