## Appendix for "Novel Application of a Multistate Model to Evaluate the Opioid Use Disorder Care Cascade: A Retrospective Cohort Study"

**Appendix A. Data Source and Extraction**

The definitions and codes used to extract data are presented here.

**Appendix Table 1: Relevant ICD-9 codes** **for Diagnosis of OUD.**

| **ICD-9-CM Code** | **ICD-9-CM Description** |
| --- | --- |
| 304.00-304.02 | Opioid type dependence (unspecified; continuous; episodic) |
| 304.70-304.72 | Combinations of opioid type drug with any other drug dependence (unspecified; continuous; episodic) |
| 305.50-305.52 | Opioid abuse (unspecified; continuous; episodic) |

ICD-9-CM=International Classification of Diseases, Ninth Revision, Clinical Modification

**Appendix Table 2: Relevant ICD-10 codes for Diagnosis of OUD.**

| **ICD-10-CM Code** | **ICD-10-CM Description** |
| --- | --- |
| F11.10 | Opioid abuse, uncomplicated |
| F11.120 | Opioid abuse with intoxication, uncomplicated |
| F11.121 | Opioid abuse with intoxication delirium |
| F11.122 | Opioid abuse with intoxication with perceptual disturbance |
| F11.129 | Opioid abuse with intoxication, unspecified |
| F11.14 | Opioid abuse with opioid-induced mood disorder |
| F11.150 | Opioid abuse with opioid-induced psychotic disorder with delusions |
| F11.151 | Opioid abuse with opioid-induced psychotic disorder with hallucinations |
| F11.159 | Opioid abuse with opioid-induced psychotic disorder, unspecified |
| F11.181 | Opioid abuse with opioid-induced sexual dysfunction |
| F11.182 | Opioid abuse with opioid-induced sleep disorder |
| F11.188 | Opioid abuse with other opioid-induced disorder |
| F11.19 | Opioid abuse with unspecified opioid-induced disorder |
| F11.20 | Opioid dependence, uncomplicated |
| F11.220 | Opioid dependence with intoxication, uncomplicated |
| F11.221 | Opioid dependence with intoxication delirium |
| F11.222 | Opioid dependence with intoxication with perceptual disturbance |
| F11.229 | Opioid dependence with intoxication, unspecified |
| F11.23 | Opioid dependence with withdrawal |
| F11.24 | Opioid dependence with opioid-induced mood disorder |
| F11.250 | Opioid dependence with opioid-induced psychotic disorder with delusions |
| F11.251 | Opioid dependence with opioid-induced psychotic disorder with hallucinations |
| F11.259 | Opioid dependence with opioid-induced psychotic disorder, unspecified |
| F11.281 | Opioid dependence with opioid-induced sexual dysfunction |
| F11.282 | Opioid dependence with opioid-induced sleep disorder |
| F11.288 | Opioid dependence with other opioid-induced disorder |
| F11.29 | Opioid dependence with unspecified opioid-induced disorder |
| F11.90 | Opioid use, unspecified, uncomplicated |
| F11.920 | Opioid use, unspecified, with intoxication, uncomplicated |
| F11.921 | Opioid use, unspecified, with intoxication delirium |
| F11.922 | Opioid use, unspecified, with intoxication with perceptual disturbance |
| F11.929 | Opioid use, unspecified, with intoxication, unspecified |
| F11.93 | Opioid use, unspecified with withdrawal |
| F11.94 | Opioid use, unspecified with opioid-induced mood disorder |
| F11.950 | Opioid use, unspecified with opioid-induced psychotic disorder with delusions |
| F11.951 | Opioid use, unspecified with opioid-induced psychotic disorder with hallucinations |
| F11.959 | Opioid use, unspecified with opioid-induced psychotic disorder, unspecified |
| F11.981 | Opioid use, unspecified with opioid-induced sexual dysfunction |
| F11.982 | Opioid use, unspecified with opioid-induced sleep disorder |
| F11.988 | Opioid use, unspecified with other opioid-induced |
| F11.99 | Opioid use, unspecified with unspecified opioid-induced disorder |

**Appendix Table 3. Current Procedural Terminology (CPT) for OUD Medications.**

| MOUD | CPT code | In-office dispensing of |
| --- | --- | --- |
| Methadone | H0020 | Methadone service |
| Buprenorphine | J0570-J0575 | Buprenorphine services |
|  | J2315 | ER injection |
| Naltrexone | Q9991-Q9992 | Injectable |

**Appendix Table 4. National Drug Codes Used to Identify Pharmacy Claims for OUD Medications. Adapted from (1, 2).**

**Buprenorphine:**

149075701, 12496075701, 12496075705, 54569141600, 54569141601, 54017613, 93537856, 228315603, 378092393, 12496127802, 35356055530, 49999063830, 50383092493, 68308020230, 54017713, 93537956, 228315303, 378092493, 12496131002, 35356055630, 49999063930, 50383093093, 63874117303, 68308020830, 74201201, 74201232, 409201232, 21695051510, 38779088800, 38779088801, 38779088803, 38779088806, 38779088809, 49452129201, 49452129202, 49452129203, 49452825301, 49452825302, 49452825303, 51552076501, 51552076502, 51552076505, 51552076506, 51552076509, 51552076510, 51552076550, 51927101200, 62991158301, 62991158302, 62991158303, 62991158304, 62991158306, 62991158307, 62991158308, 63275992201, 63275992202, 63275992203, 63275992204, 63275992205, 63275992207, 63370090506, 63370090509, 63370090510, 63370090515, 54018813, 93572056, 228315403, 228315473, 406192303, 12496128302, 16590066630, 42291017430, 49999039507, 49999039515, 49999039530, 50383029493, 52959074930, 54569549600, 54868575000, 55700018430, 63629402801, 63874108503, 65162041603, 68071151003, 68258299903, 54018913, 93572156, 228315503, 228315573, 406192403, 12496130602, 35356000407, 35356000430, 42291017530, 43063018407, 43063018430, 50383028793, 52959030430, 54569573900, 54569573901, 54569573902, 54569640800, 54868570700, 54868570701, 54868570702, 54868570703, 54868570704, 55045378403, 63629403401, 63629403402, 63629403403, 63874108403, 65162041503, 66336001630, 68071138003, 40042001001, 42023017901, 42023017905, 55390010010, 35356060704, 59011075004, 35356060504, 54569632500, 59011075104, 35356060604, 54569632600, 59011075204, 12496120201, 12496120203, 12496120801, 12496120803, 54569639900, 55700014730, 12496120401, 12496120403, 12496121201, 12496121203, 54123091430, 54123095730, 59011075804, 59385001201, 59385001230, 59385001401, 59385001430, 59385001601, 59385001630, 59011075704, 54123098630, 54123011430, 54123092930, 63481016101, 63481016160, 63481020701, 63481020760, 63481034801, 63481034860, 63481051901, 63481051960, 63481068501, 63481068560, 63481082001, 63481082060, 63481095201, 63481095260, , 143924605, 378876593, 378876693, 378876793, 378876893, 406800503, 406802003, 517072505, 781721664, 781722764, 781723864, 781724964, 12496010001, 12496030001, 42858050103, 42858050203, 42858060103, 42858060203, 43598057930, 43598058030, 43598058130, 43598058230, 47781035503, 47781035603, 47781035703, 47781035803, 50268014415, 50268014515, 52427069203, 52427069403, 52427069803, 52427071203, 52440010014, 54123090730, 55700030230, 58118017608, 58118017708, 58118315608, 60429058630, 60429058730, 60687048121, 60687049221, 62175045232, 62175045832, 62756045964, 62756045983, 62756046064, 62756046083, 62756096964, 62756096983, 62756097064, 62756097083, 63629712601, 63629712602, 63629712603, 63629712604, 63629712605, 63629712606, 63629712607, 63629712608, 63629712609, 65162041509, 65162041609, 67046099430, 67046099530, 67046099630, 67046099730, 70518071100, 70518071101, 70518071102, 70518100700, 70518155700, 70518162500, 70518168400, 70518201400, 70518221600, 70518221700, 70518221800, 70518222600, 70518231100, 70518232700, 71335035301, 71335035302, 71335035303, 71335035304, 71335035305, 71335035306, 71335035307, 71335095001, 71335095002, 71335095003, 71335095004, 71335095005, 71335095006, 71335095007, 71335115401, 71335115402, 71335115403, 71335115404, 71335115405, 71335115406, 71335115407, 71335115408, 71335115409, 71335116301, 71335116302, 71335116303, 71335116304, 71335116305, 71335116306, 71335116307, 71335116308, 71335116309, 71335129601, 71335129602, 71335137801, 71335151401, 71335151402, 71335165301

**Naltrexone:**

56001122, 56001130, 56001170, 56007950, 56008050, 185003901, 185003930, 406009201, 406009203, 406117001, 406117003, 555090201, 555090202, 16729008101, 16729008110, 42291063230, 43063059115, 47335032683, 47335032688, 50436010501, 51224020630, 51224020650, 51285027501, 51285027502, 52152010502, 52152010504, 52152010530, 54868557400, 65694010003, 65694010010, 68084029111, 68084029121, 68094085362, 68115068030, 49452483501, 51927275300, 51927360200, 51927354800, 63459030042, 65757030001, 65757030202, 38779088703, 38779088704, 38779088705, 38779088706, 38779088708, 49452480801, 49452480802, 49452483502, 51552073701, 51552073702, 51552073704, 51927437700, 52372075101, 52372075102, 52372075103, 55812033301, 55812033302, 55812033303, 63370015810, 63370015815, 63370015825, 63370015835, 63370015845, 60966014404, 60966024403, 60966034402, 62991124301, 62991124302, 62991124303, 62991124304

**Appendix Table 5. Current Procedural Terminology (CPT) and Healthcare Common Procedure Coding System (HCPCS) Codes Used to Identify Utilization of Behavioral health services (i.e., mental health or substance use disorder treatment). Adapted from (3).**

**Behavioral health services in any setting:**

90791-90792, 90832-90834, 90836-90840, 90853, 90863, 90875-90876, G0396, G0397, H0001, H0002, H0004, H0005, H0007-H0011, H0012, H0013, H0014, H0015, H0016, H0019, H0022, H0031, H0034-H0037, H0039, H0040, H0047, H2000, H2001, H2010-H2020, H2035, H2036, M0064, S0201, S9480, S9484, S9485, T1006, T1012

Among above codes, those indicating detoxification services: H0008-H0014

Among above codes, those indicating intensive outpatient care: H0015

Among above codes, those indicating partial hospitalization: H0035

Among above codes, those indicating residential care: H0012, H0013, H0017-H0019

Among above codes, those indicating inpatient care: H0008-H0011

**Appendix Table 6: Relevant ICD-9-CM and ICD-10-CM Codes for Clinical and Behavioral Variables. Adapted from (4).**

| **Variables** | **ICD-9-CM/ICD-10-CM Code** |
| --- | --- |
| **Substance Use** | |
| Marijuana | 30430, 30431, 30432, F1210, F1212, F1215, F1218, F1219, F1220, F1222, F1225, F1228, F1229, F1290, F1292, F1295, F1298, F1299 |
| Cocaine | 30420, 30421, 30422, 30560, 30561, 30562, F1410, F1412, F1414, F1415, F1418, F1419, F1420, F1422, F1424, F1425, F1428, F1429, F1490, F1492, F1494, F1495, F1498, F1499 |
| Tobacco | 3051, 30510, F1720, F1721, F1722, F1729, T6522, T6529, Z720 |
| Alcohol | 2911, 2912, 2913, 2914, 2915, 2918, 29180, 29181, 29182, 29189, 30300, 30301, 30302, 30390, 30391, 30392, 30500, 30501, 30502, 53530, 53531, F1010, F1012, F1015, F1018, F1019, F1020, F1022, F1023, F1024, F1025, F1028, F1029, F1092, F1094, F1095, K2920, K2921 |
| **Overdose** | |
|  | 96500, 96501, 96502, 96509, E8500, E8501, E8502, T400, T401, T402, T403 |
| **Pain** | |
| Lower back pain | 7245, 7248, 72458, 7249, M545, M546, M5489, M549, M6283 |
| Neck pain | 52461, 52462, 52463, 52464, 52469, M2660, M2661, M2662, M2663 |
| Headache | 30781, 34600, 34601, 34602, 34603, 34610, 34611, 34612, 34613, 34620, 34621, 34622, 34623, 34630, 34631, 34632, 34633, 34650, 34651, 34652, 34653, 34661, 34670, 34671, 34672, 34673, 34680, 34681, 34682. 34683, 34690, 34691, 34692, 34693, 7840, G4300, G4301, G4310, G4311, G4340, G4341, G4350, G4351, G4370, G4371, G4380, G4381, G4382, G4383, G4390, G4391, G43A0, G43A1, G43B0, G43B1, G43C0, G43C1, G43D0, G43D1, G4400, G4401, G4402, G4403, G4404, G4405, G4409, G441, G4420, G4421, G4422, G4430, G4431, G4432, G4440, G4441, G4451, G4452, G4453, G4459, G4481, G4482, G4483, G4484, G4485, G4489, R51 |
| Fibromyalgia | 725, M353, M7910, M7911, M7912, M7918, M797, R5382 |
| Osteoarthritis | 71516, 71517, 71518, 71520, 71521, 71522, 71523, 71524, 71525, 71526, 71527, 71528, 71530, 71531, 71532, 71533, 71534, 71535, 71536, 71537, 71580, 71589, M150, M151, M152, M153, M154, M158, M159, M160, M1610, M1611, M1612, M162, M1630, M1631, M1632, M164, M1650, M1651, M1652, M166, M167, M169, M170, M1710, M1711, M1712, M1730, M1731, M1732, M174, M175, M179, M180, M1810, M1811, M182, M1830, M1831, M179, M180, M1810, M1811, M1812, M182, M1830, M1831, M1832, M184, M1850, M1851, M1852, M189, M1901, M1902, M1903, M1904, M1907, M1911, M1912, M1913, M1914, M1917, M1921, M1922, M1923, M1924, M1927, M1990, M1991, M1992, M1993 |
| Rheumatoid arthritis | 7140, 7141, 7142, 71430, 71431, 71432, M0510, M0514, M0516, M0519, M0520, M0523, M0524, M0529, M0530, M0531, M0540, M0543, M0544, M0545, M0546, M0547, M0549, M0550, M0553, M0554, M0555, M0556, M0559, M0560, M0561, M0562, M0564, M0566, M0567, M0569, M0570, M0571, M0572, M0573, M0574, M0575, M0576, M0577, M0579, M0580, M0581, M0582, M0583, M0584, M0585, M0586, M0587, M0589, M059, M0600, M0601, M0602, M0603, M0604, M0605, M0606, M0607, M0609, M061, M0621, M0622, M0623, M0625, M0627, M0628, M0630, M0632, M0634, M0637, M0639, M064, M0680, M0681, M0682, M0683, M0684, M0685, M0686, M0687, M0688, M0689, M069, M0800, M0801, M0803, M0804, M0807, M0808, M0809, M081, M0820, M0823, M0824, M0829, M083, M0840, M0843, M0844, M0848, M0880, M0883, M0890, M0899 |
| Sprain | 8400, 8401, 8402, 8403, 8404, 8405, 8406, 8407, 8408, 8409, 8410, 8411, 8412, 8413, 8418, 8419, 84200, 84201, 84202, 84209, 84210, 84211, 84212, 84213, 84219, 8430, 8431, 8438, 8439, 8440, 8441, 8442, 8443, 8448, 8449, 84500, 84501, 84502, 84503, 84509, 84510, 84511, 84512, 84513, 84519, 8460, 8461, 8462, 8468, 8469, 8470, 8472, 8473, 8479, 8480, 8481, 8482, 8483, 84840, 84841, 84842, 84849, 8485, 8488, 8489, S031X, S0340, S0341, S0342, S034X, S039X, S0911, S134X, S135X, S138X, S139X, S161X, S233X, S2341, S2342, S238X, S239X, S2901, S335X, S336X, S338X, S339X, S3901, S4340, S4341, S4342, S4343, S4349, S4350, S4351, S4352, S4361, S4362, S4380, S4381, S4382, S4390, S4391, S4392, S4601, S4611, S4621, S4631, S4681, S4691, S5320, S5321, S5322, S5330, S5331, S5332, S5340, S5341, S5342, S5343, S5344, S5349, S5601, S5611, S5621, S5631, S5641, S5651, S5681, S5691, S6330, S6331, S6332, S6339, S6340, S6341, S6343, S6349, S6350, S6351, S6352, S6359, S6360, S6361, S6362, S6363, S6364, S6365, S6368, S6369, S638X, S6390, S6391, S6392, S6601, S6611, S6621, S6631, S6641, S6651, S6681, S6691, S7310, S7311, S7312, S7319, S7601, S7611, S7621, S7631, S7681, S7691, S8340, S8341, S8342, S8350, S8351, S8352, S8360, S8361, S8362, S838X, S8390, S8391, S8392, S8601, S8611, S8621, S8631, S8681, S8691, S9340, S9341, S9342, S9343, S9343, S9349, S9350, S9351, S9352, S9360, S9361, S9362, S9369, S9601, S9611, S9621, S9681, S9691 |
| **Clinical Disorders** | |
| Adjustment Disorder | 3090, 3091, 30922, 30923, 30924, 30928, 30929, 3093, 3094, 30982, 30983, 30989, 3099, F4320, F4321, F4322, F4323, F4324, F4325, F4329, F438, F439 |
| Mood Disorder | 29383, 29600, 29601, 29602, 29603, 29604, 29605, 29606, 29610, 29611, 29612, 29613, 29614, 29615, 29616, 29620, 29621, 29622, 29623, 29624, 29625m 29626, 29630, 29631, 29632, 29633, 29634, 29635, 29636, 29640, 29641, 29642, 29643, 29644, 29645, 29646, 29650, 29651, 29652, 29653m 29654, 29655, 20656, 29660, 29661, 29662, 29963, 29664, 29665, 29666, 2967, 29670, 29680, 29681, 29682, 29689, 29690, 29699, 3004, 311, 3110, 31119, F0630, F0631, F0632, F0633, F0634, F3010, F3011, F3012, F3013, F302, F303, F304, F308, F309, F310, F3110, F3111, F3112, F3113, F312, F3130, F3131, F3132, F314, F315, F3160, F3161, F3162, F3163, F3164, F3170, F3171, F3172, F3173, F3174, F3175, F3176, F3177, F3178, F3181, F319, F320, F321, F322, F323, F324, F325, F328, F3281, F3289, F329, F330, F331, F332, F333, F3340, F3341, F3342, F338, F339, F340, F341, F348, F3481, F3489, F349, F39, R4586 |
| Personality Disorder | 3010, 30110, 30112, 30113, 30120, 30121, 30122, 3013, 3014, 30150, 30151, 3016, 3017, 30181, 30182, 30183, 30184, 30189, 3019, F600, F601, F602, F603, F604, F605, F606, F607, F6081, F6089, F609, F69 |
| Anxiety Disorder | 29384, 30000, 30001, 30002, 30009, 30010, 30020, 30021, 30022, 30023, 30029, 3003, 3005, 30089, 3009, 3080, 3081, 3082, 3083, 3084, 3089, 30981, 3130, 3131, 31321, 3133, F064, F4000, F4001, F4002, F4010, 4011, F4021, F4023, F4024, F4029, F408, F409, F410, F411, F413, F418, F419, F42, F422, F423, F424, F428, F429, F430, F4310, F4311, F4312, F488, F489, R452, R453, R454, R455, R456, R457, R4581, R4582, R4583, R4584 |
| **Other Clinical Factors** | |
| HCV | Pegasys, pegintron, ribavirin, sofosbuvir, viekira, zepatier |
| HIV | 042, 043, 044, B20, B21, B22, B24 |
| CCI_b | Includes myocardial infarction, congestive heart failure, peripheral vascular disease, cerebrovascular disease, dementia, chronic pulmonary disease, connective tissue disease-rheumatic disease, peptic ulcer disease, mild liver disease, diabetes without complications, diabetes with complications, paraplegia and hemiplegia, renal disease, cancer, moderate or severe liver disease, metastatic carcinoma |

**Appendix Figure 1: Flow diagram describing the study sample selection process.**

OUD during January 2012 to March 2019 defined as:

- admissions/primary/secondary dx from inpatient
- or primary dx from outpatient
- or secondary dx from outpatient ED setting

n=12,249

Patients that had been continuously enrolled during

baseline (one-year before index OUD dx) and more

than 30 days of follow-up

n=6,939

Adult patients had OUD dx before February 2018

n=10,840

Adult patients had OUD dx

n=12,173

Excluded since OUD dx criteria not met

n=76

Excluded since OUD dx criteria was not before February 2018

n=1,333

Excluded since were not enrolled continuously during baseline

n=3,901

**Appendix B. Multistate Model Definition and Assumptions Assessment**

We briefly describe the model described in detail in (5, 6) here. Let $X_{ij}$ denote the covariate matrix for person $i$at time $j$, $S_{ij}$ denotes state membership for person $i$at time $j,$ $L$ is the total number of states, $p_{jkl}=P(S_{j}=l|S_{j-1}=k)$ is the state transition probability, $t_{j}$is a calendar date corresponding to time interval, and $h_{kl}\left( d_{ij};\gamma_{kl} \right)$is the transition-specific period effect. Let $p_{jkl}=pr(S_{j}=l |S_{j-1}=k)$ represent the state transition probability, *P_j_* for a given state *S* at time *j* given the previous state *j-1*. The estimated transition matrix is denoted **P**_j_. A multistate model can take the form $log\left( \frac{p_{jkl}\left( x_{ij},d_{ij} \right)}{p_{jk1}\left( x_{ij},d_{ij} \right)} \right)=\alpha_{jkl}+x_{j}\beta_{kl}+ h_{kl}(d_{ij};\gamma_{kl})$, with $k=1,...,8$ and $l=2,...,8$.

We assessed lack of fit for the multistate model by creating plots of observed versus fitted state membership probabilities for each time interval. In Appendix Figure 2, the predicted proportions were calculated using the multistate model with covariates to estimate predicted state membership probabilities at each time point. The standard error of the predicted proportion in each state were estimated using bootstrap resampling with 200 bootstrap replicates (7). The points overlap for all states over time except the initial time intervals in RE1 and RE2. We evaluated the second order dependence by also including second order state membership at $t_{j-2}$ as a categorical covariate. In Appendix Figure 3, we plot the observed and predicted state probabilities with the second order state membership, $S_{j-2}$ , as a covariate in the model. These combined results demonstrated that for most states, the first order Markov assumption is not violated and there was no indication of a lack of model fit (5, 6).

In Appendix Figure 4 – 6, we examined the relative risk ratios when the multistate model also included the second order state membership. Comparing these results in Appendix Figure 4 – 6 with the main document Figures 2-4, we observed that inclusion of second order state membership did not meaningfully change our findings. Thus, the association of covariates with transition rates estimated in the two multistate models appears to be fairly robust to this assumption of first or second order Markov dependence.

**Appendix Figure 2: Model assumption assessment by evaluating observed and predicted marginal state probabilities.**

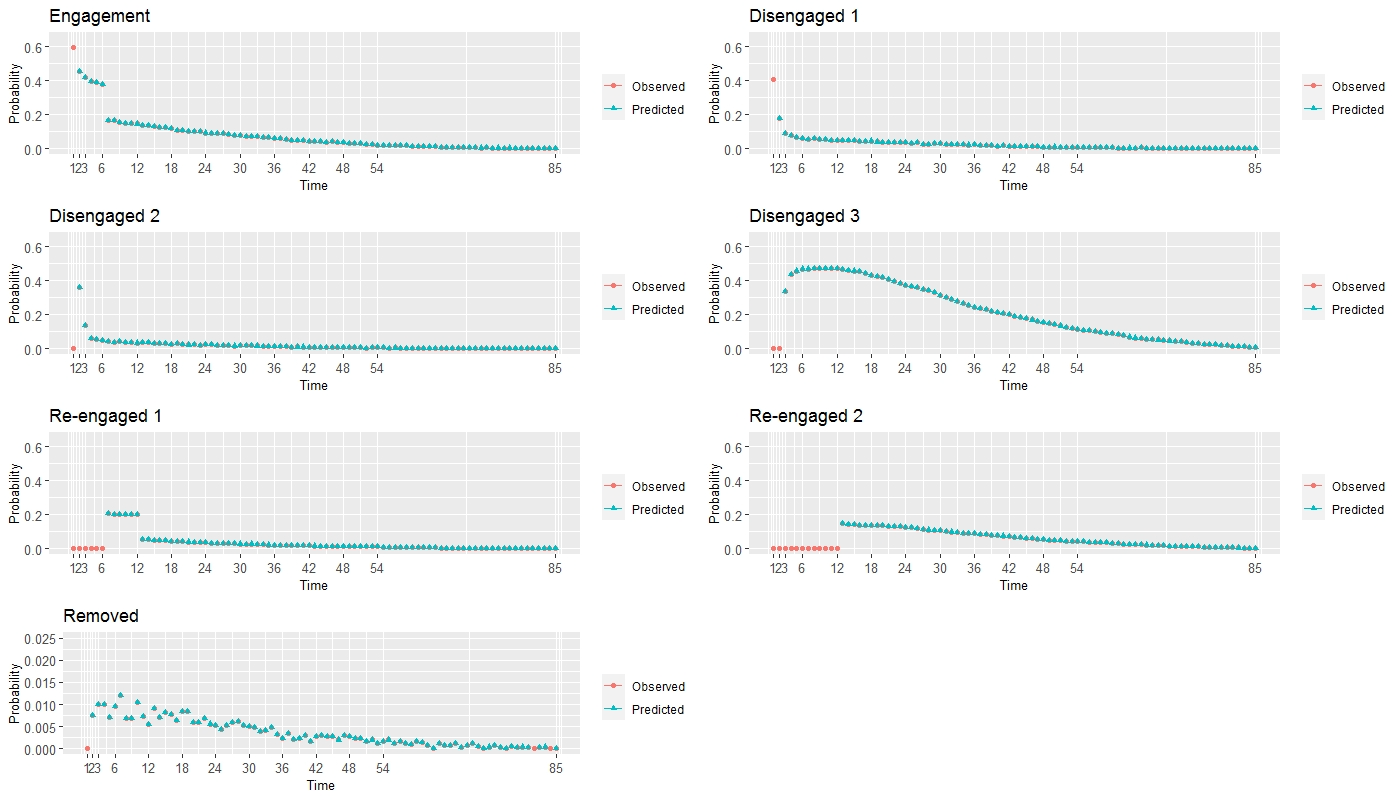

**Appendix Figure 3: Model assumption assessment by evaluating observed and predicted marginal state probabilities.**

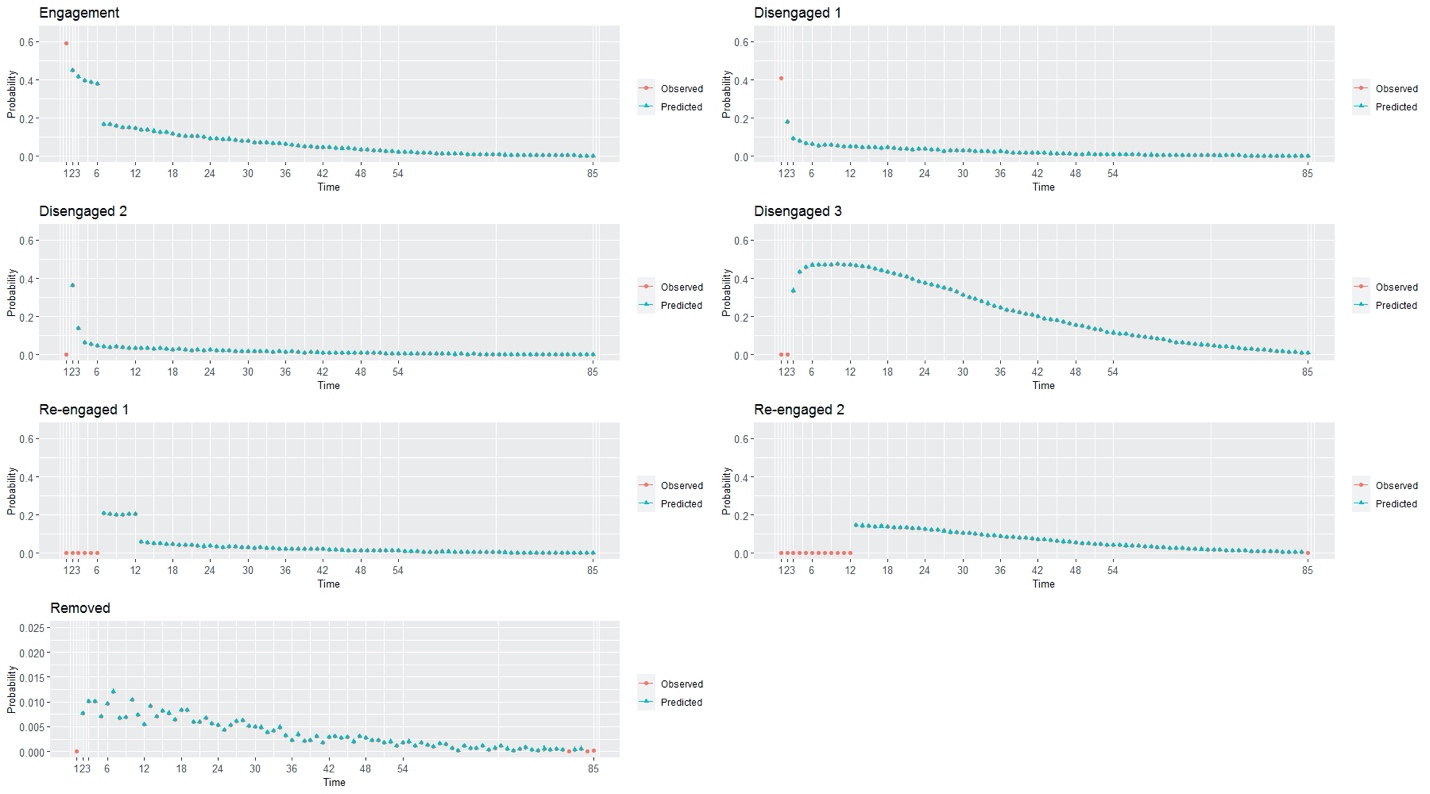

**Appendix Figure 4: Model assumptions evaluated by assessing the relative risk ratios^a^ capturing transitions from engagement to early retention (EN to RE1, panel A) and from engagement to early discontinuation (EN to DS1, panel** **B) among patients with behavior and/or medication therapy.**

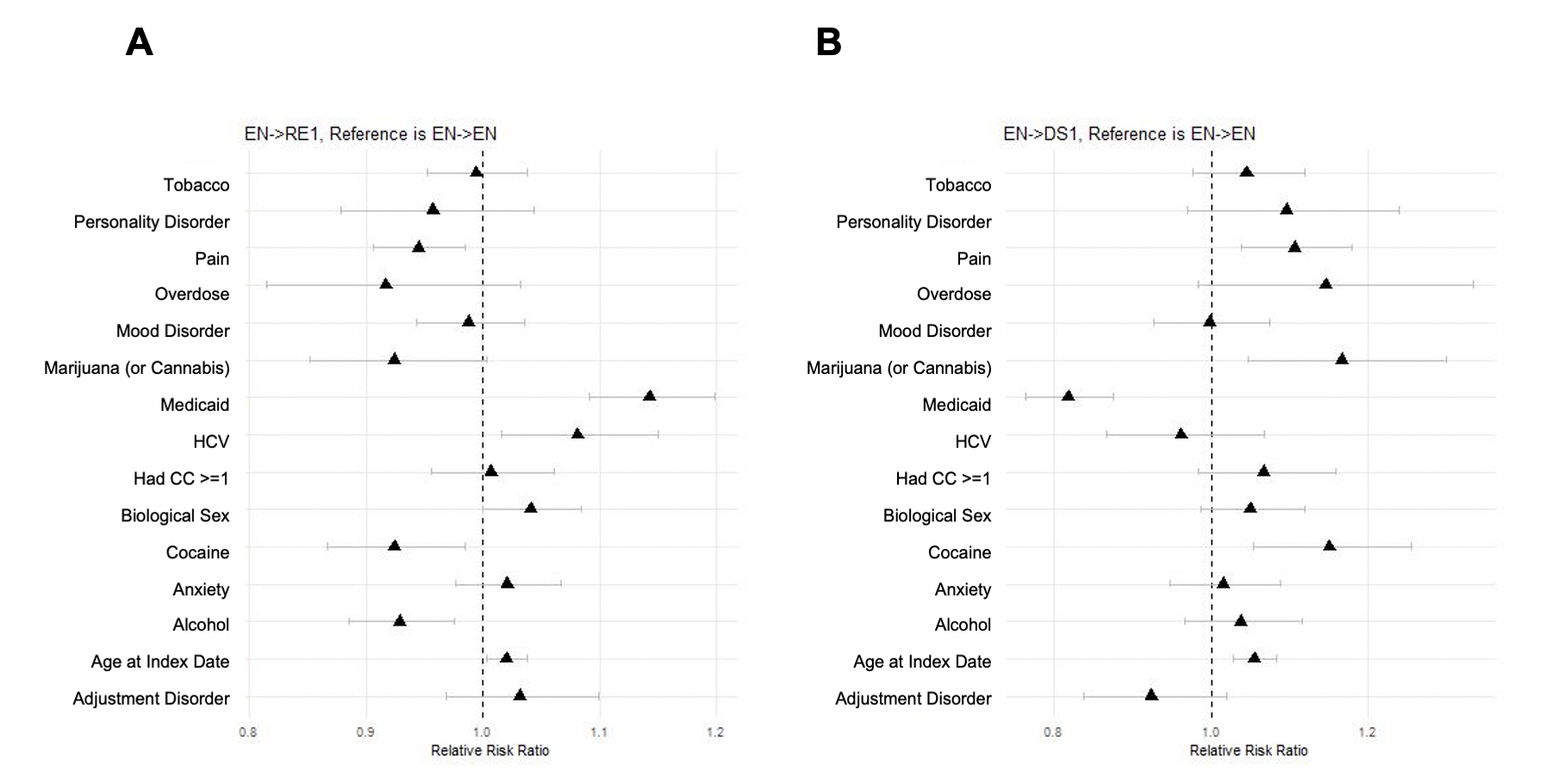

^a^ Reference group for variables defined as follows: “other (commercial or Medicare)” for Medicaid, “male” for biological sex, and “no” for tobacco use disorder, personality disorder, pain, overdose, mood disorder, marijuana (or cannabis), HCV, had CC >= 1, cocaine use disorder, anxiety disorder, alcohol use disorder, and adjustment disorder. Age at index date measured in years.

**Appendix Figure 5: Model assumptions evaluated by assessing the relative risk ratios^a^ capturing transitions from short- and long-term retention. RE1 to RE2 in (panel A), RE1 to DS1 in (panel B), and RE2 to DS1 in (panel C) among patients with behavior and/or medication therapy.**

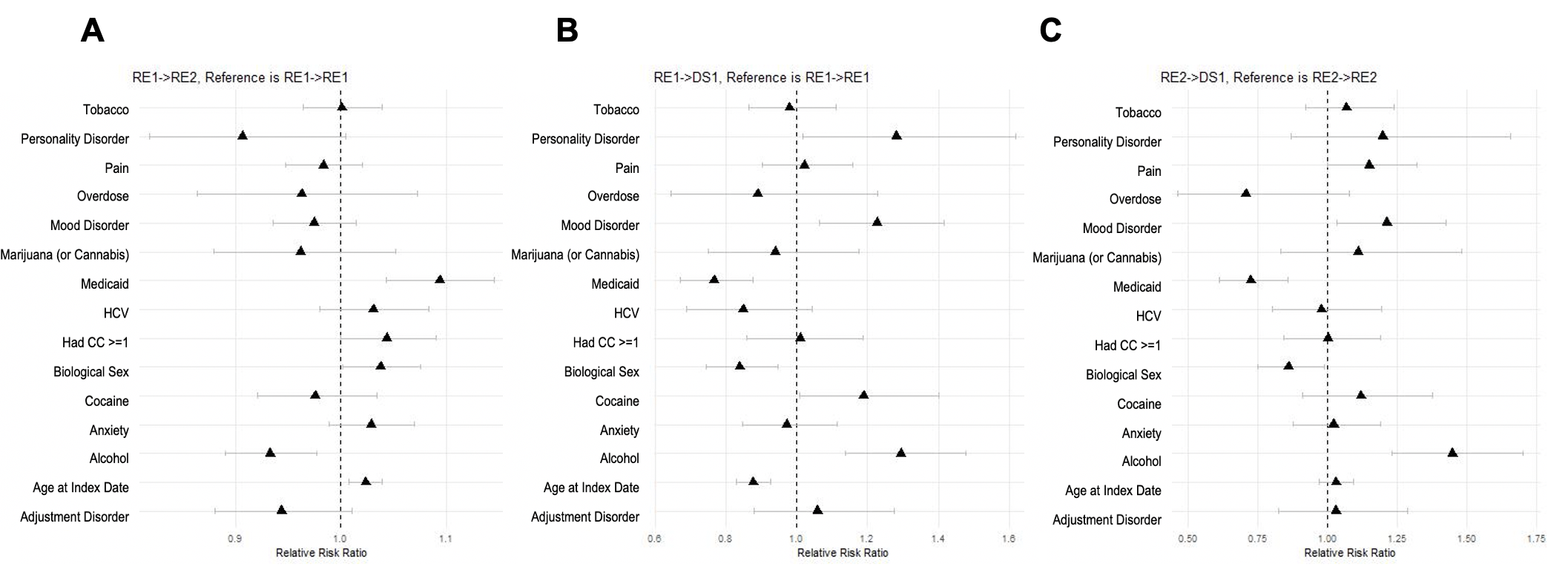

^a^ Reference group for variables defined as follows: “other (commercial or Medicare)” for Medicaid, “male” for biological sex, and “no” for tobacco use disorder, personality disorder, pain, overdose, mood disorder, marijuana (or cannabis), HCV, had CC >= 1, cocaine use disorder, anxiety disorder, alcohol use disorder, and adjustment disorder. Age at index date measured in years.

**Appendix Figure 6: Model assumptions evaluated by assessing the relative risk ratios^a^ capturing transitions from short- and long-term disengagement. DS1 to EN in (panel A), DS2 to EN in (panel B), and DS3 to EN in (panel C) among patients with behavior and/or medication therapy.**

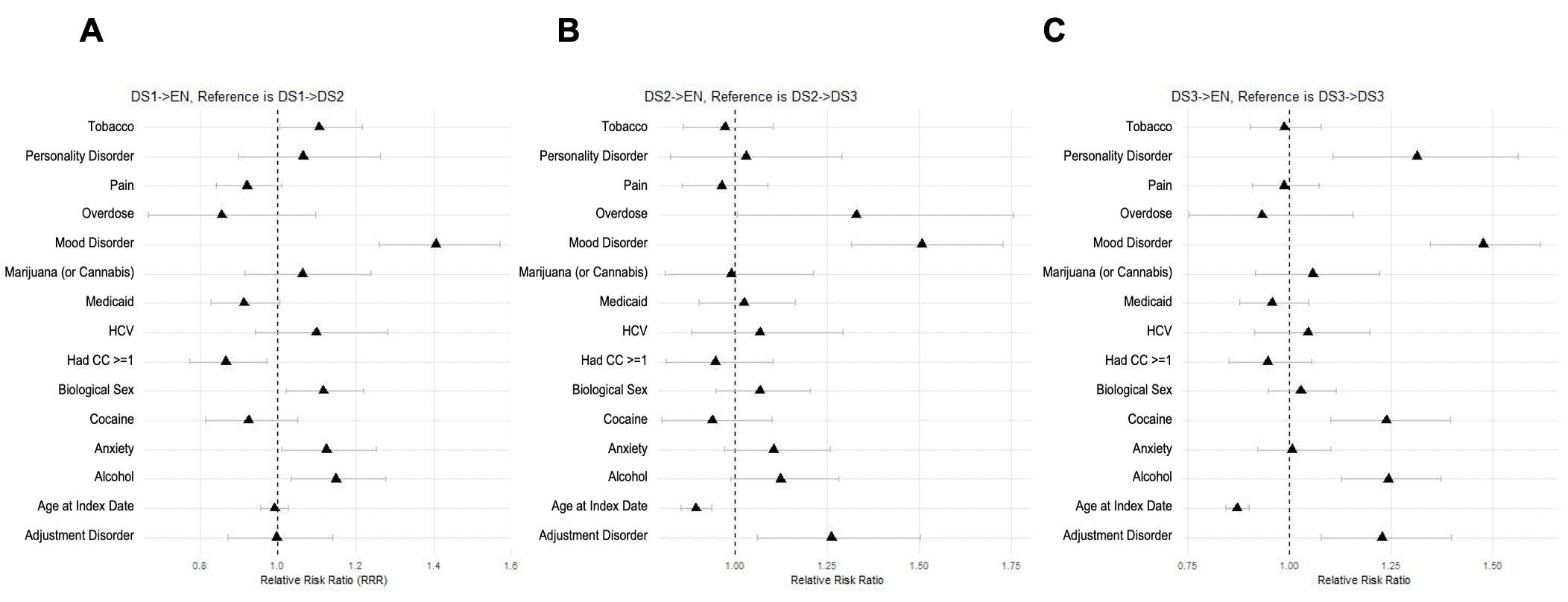

^a^ Reference group for variables defined as follows: “other (commercial or Medicare)” for Medicaid, “male” for biological sex, and “no” for tobacco use disorder, personality disorder, pain, overdose, mood disorder, marijuana (or cannabis), HCV, had CC >= 1, cocaine use disorder, anxiety disorder, alcohol use disorder, and adjustment disorder. Age at index date measured in years.

**C. Sensitivity Analysis**

This section contains results of the sensitivity analysis. The primary analysis defined OUD treatment as either behavioral and/or medication therapy. We conducted a sensitivity analysis classifying OUD treatment as medication only. The relative risk ratios (RRR) with 95% confidence intervals are shown below for the medication (M) only for each sub-stage below.

**Appendix Figure 7: Relative risk ratios^a^ capturing engagement, EN to RE1, in (A) and early disengagement, EN to DS1, in (B). The M model refers to the results in which medication treatment only and BM model refers to the results in which medication and/or behavioral therapy are included.**

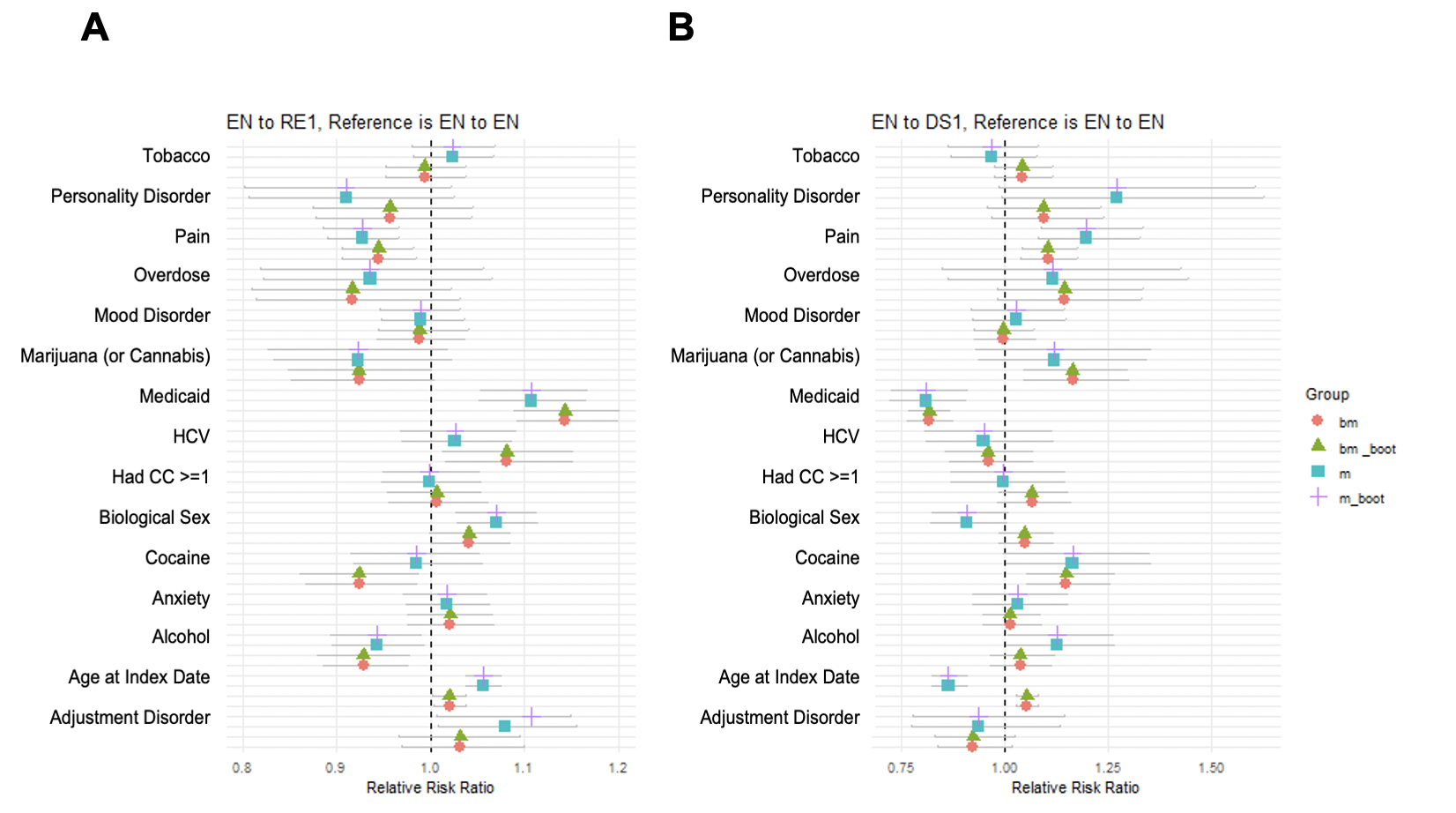

^a^ Reference group for variables defined as follows: “other (commercial or Medicare)” for Medicaid, “male” for biological sex, and “no” for tobacco use disorder, personality disorder, pain, overdose, mood disorder, marijuana (or cannabis), HCV, had CC >= 1, cocaine use disorder, anxiety disorder, alcohol use disorder, and adjustment disorder. Age at index date measured in years.

**Appendix Figure 8: Relative risk ratios^a^ capturing Short- and Long-Term Disengagement. DS1 to EN in (A), DS2 to EN in (B), and DS3 to EN in (C). The M model refers to the results in which medication treatment only and BM model refers to the results in which medication and/or behavioral therapy are included.**

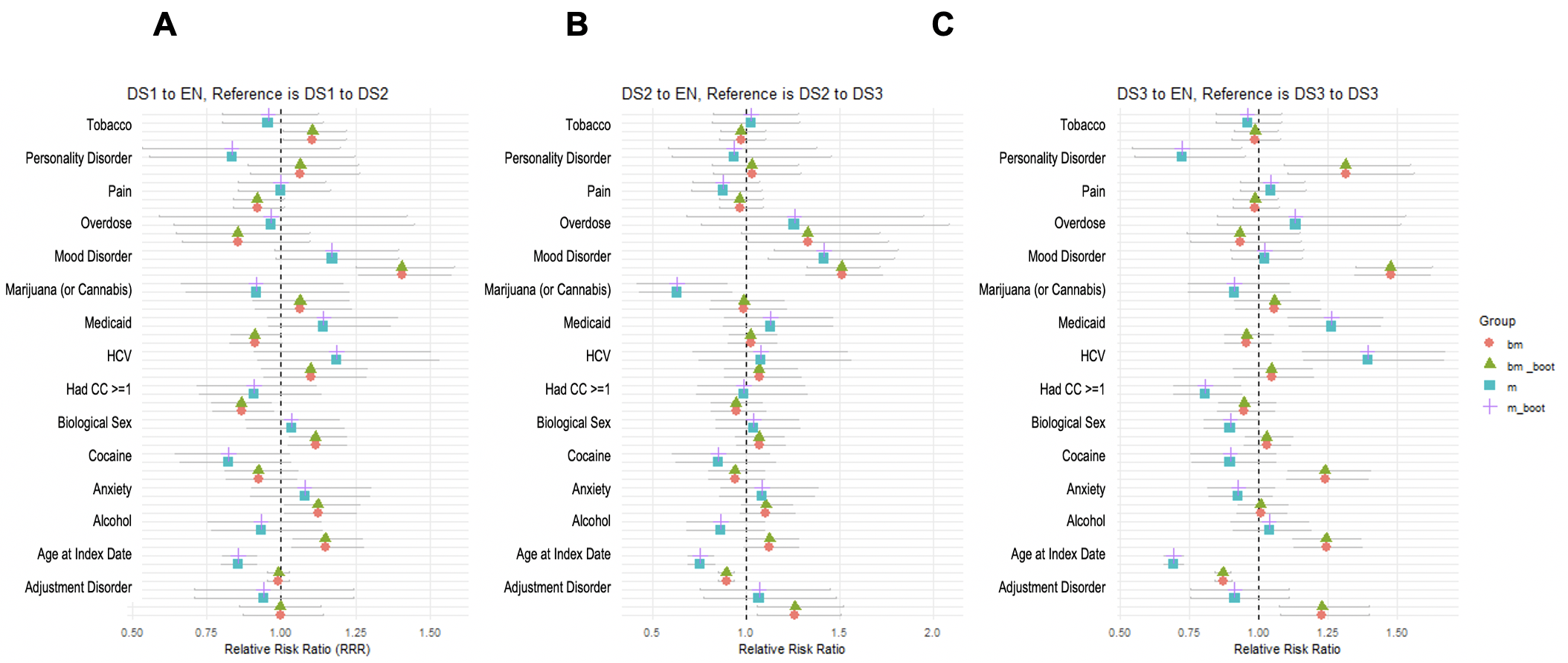

^a^ Reference group for variables defined as follows: “other (commercial or Medicare)” for Medicaid, “male” for biological sex, and “no” for tobacco use disorder, personality disorder, pain, overdose, mood disorder, marijuana (or cannabis), HCV, had CC >= 1, cocaine use disorder, anxiety disorder, alcohol use disorder, and adjustment disorder. Age at index date measured in years.

**Appendix Figure 9: Relative risk ratios^a^ capturing Short- and Long-Term Retention. RE1 to RE2 in (A), RE1 to DS1 in (B), and RE2 to DS1 in (C). The M model refers to the results in which medication treatment only and BM model refers to the results in which medication and/or behavioral therapy are included.**

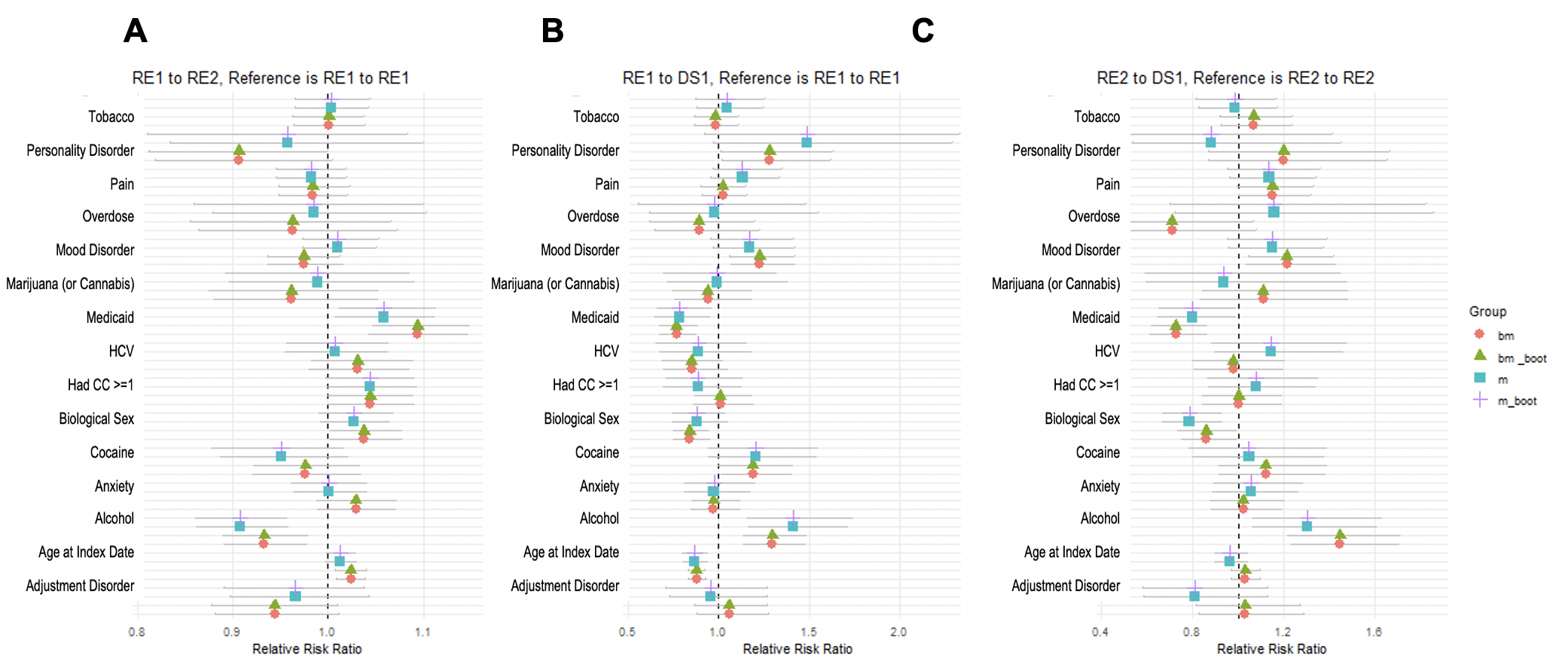

^a^ Reference group for variables defined as follows: “other (commercial or Medicare)” for Medicaid, “male” for biological sex, and “no” for tobacco use disorder, personality disorder, pain, overdose, mood disorder, marijuana (or cannabis), HCV, had CC >= 1, cocaine use disorder, anxiety disorder, alcohol use disorder, and adjustment disorder. Age at index date measured in years.

**D. Additional Results**

This section contains additional results not included in the main document. A detailed description of definitions used to classify patients into each state using claims data is summarized in Appendix D Table 7. Estimated transition probabilities between states are shown in Appendix D Table 8 and estimates over time shown in Appendix D Figure 7.

**Appendix Table 7. Definition of States in the Multistate Modeling Framework.**

| **state** | **Description** | APCD Definition |
| --- | --- | --- |
| Diagnosed with OUD | Persons identified with OUD diagnosis (ICD-9/10-CM code) | Inpatient admission had OUD as primary or secondary diagnosis.  Emergency department visit with OUD as primary or secondary diagnosis.  Outpatient visit with OUD as primary diagnosis to have high specificity. |
| Engaged | If MOUD or behavioral therapy services <= 180 days | At least one medical encounter for behavioral therapy, at least one CPT code for methadone, or a seven-day supply of OUD medication within one month of the initial OUD |
| Retained state I | If MOUD or behavioral therapy services 180 to 360 days | Consecutive intervals on treatment between 180 to 360 days after engaging in treatment. |
| Retained state 2 | If MOUD or behavioral therapy services > 360 days | Consecutive intervals on treatment for more than 360 days after engaging in treatment. |
| Disengaged state 1 | Did not start or no treatment for 30 days | Treatment was missed at least once within 30 days after initial OUD diagnosis. |
| Disengaged state 2 | No treatment for 60 days | Treatment was missed at least once within 60 days after initial OUD diagnosis. |
| Disengaged state 3 | No treatment for more than 90 days | Treatment was missed once within 90 days after initial OUD diagnosis. |
| Removal | Disenrolled or deceased | Records were not present in next time intervals due to disenrollment, deceased, or other unknown reason. |

**Appendix Table 8. Probability Transition States Estimated from the Multistate Model.**

|  | **State at t_j_** | | | | | | |
| --- | --- | --- | --- | --- | --- | --- | --- |
| **State at t_j-1_** | Engaged | Retained 1 | Retained 2 | Disengaged 1 | Disengaged 2 | Disengaged 3 | Removal |
| Engaged | 0.1290 | 0.0150 | 0 | 0.0465 | 0 | 0 | 0.0012 |
| Retained 1 | 0 | 0.0560 | 0.0087 | 0.0053 | 0 | 0 | 0.0003 |
| Retained 2 | 0 | 0 | 0.1214 | 0.0045 | 0 | 0 | 0.0006 |
| Disengaged 1 | 0.0178 | 0 | 0 | 0 | 0.0477 | 0 | 0.0006 |
| Disengaged 2 | 0.0076 | 0 | 0 | 0 | 0 | 0.0387 | 0.0004 |
| Disengaged 3 | 0.0226 | 0 | 0 | 0 | 0 | 0.4700 | 0.0049 |
| Removal | 0 | 0 | 0 | 0 | 0 | 0 | 1.0000 |

**Appendix Figure 10: Estimating Transition States Over Time for Early Engagement and Retention (in A) and Short and Long-term Disengagement (in B).**

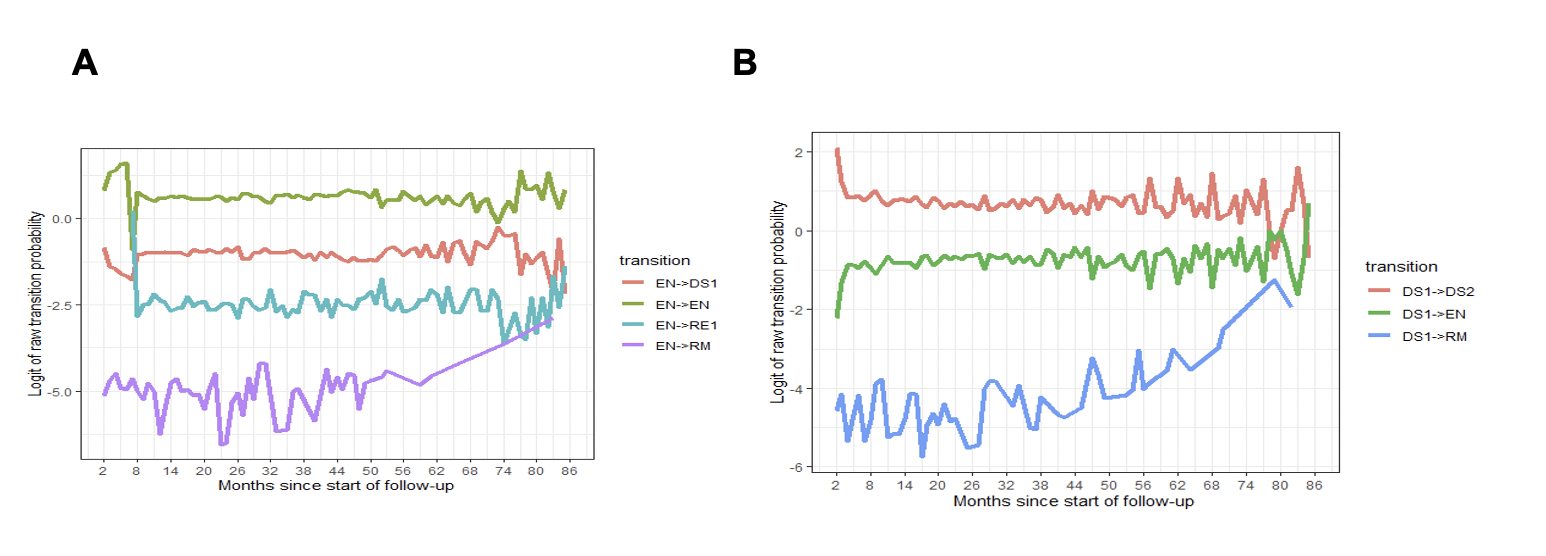
